# Discontinuation of isolation for persons with COVID-19: Is 10 days really safe?

**DOI:** 10.1101/2021.01.29.21250753

**Authors:** Alvina Clara Felix, Anderson V de Paula, Andreia Cristina Ribeiro, Francini Camila da Silva, Marta Inemami, Angela A Costa, Cibele Odete D Leal, Walter M Figueiredo, Claudio Sérgio Pannuti, Camila Malta Romano

## Abstract

**Background:** The detection of SARS-CoV-2 RNA by real-time polymerase chain reaction (PCR) in respiratory samples from COVID-19 patients is not a direct indication of the presence of viable viruses. The isolation of SARS-CoV-2 in cell culture system however, can acts as surrogate marker of infectiousness. Cell culture based studies performed mostly with hospitalized and moderate/severe COVID-19 claims that no replication competent virus is found after 9 days of the symptoms onset in respiratory samples. Therefore, it is now recommended 10 days isolation before patient discharge.

**Methods:** We cell-cultured 29 SARS-COV-2 RT-PCR positive respiratory samples at the 10^th^ day after the illness in Vero E6 cells. After two passages, cytopathic effect and cycle threshold (CT) lower than the obtained in the original sample were used to determine positivity.

**Findings:** We found viable particles in (7/29) 24% of samples tested. The positivity in cell culture was strongly associated (p<0.0001) to the low cycle thresholds in clinical samples (Ct <21).

**Conclusion:** This data adds important knowledge to the current protocols for de-isolation of patients with non-hospitalized mild COVID-19.

## Introduction

In June 2020, the Center for Diseases Control (CDC) updated the information about the time of isolation for COVID-19 patients that would prevent transmission to a susceptible individual. For exposed persons with no SARS-COV-2 confirmation, 10 days quarantine would be sufficient to reduce the transmission risk to 1% [1]. For confirmed mild and moderate COVID-19, the isolation can be also discontinued after 10 days after symptoms onset considering resolution of fever for at least 24 hours. This period however, would be extended in case of severe COVID-19, in patients presenting immunological conditions or in case the patient still reports symptoms [1].

Criteria for discharge patients without a negative RT-PCR involve risks but some issues were taken into consideration for the new resolution; in particular, the amount of accessible laboratory supplies for molecular SARS-COV-2 tests as well trained and available lab technicians in some areas. Such demand makes the initial recommendation of two negative RT-PCR tests for discharging nearly impossible to be done, especially in regions with high number of cases.

The primary recommendation of 14 days isolation for COVID-19 was based on initial description that patients with moderate/severe disease could remain RT-PCR positive until day 12-14 [2,3]. More recent studies revealed that positivity at upper and lower respiratory tract can last longer than 14 days with highest peak at day 3-5 of illness, with consistent decline over time [4,5]. However, despite viral RNA detection by RT-PCR no replication-competent virus (RCV) has been recovered from samples collected after 8 [6] or 9 days of the onset [5,7].

The time of infectiousness appears to be related to the disease severity, since accumulated evidences point that asymptomatic individuals clear the virus faster than symptomatic ones [8]. A few studies that successfully isolated SARS-COV-2 from respiratory samples also correlated the positivity in cell culture to RT-PCR cycle threshold (C_T_) values on the original clinical samples [6, 9].

Given the isolation interruption at the 10^th^ day after the illness onset is becoming widely adopted and the presence of viable particles at this time was little evaluated so far, we explored the presence of replicating-competent viruses (RCV) in RT-PCR positive nasopharyngeal (NP) samples from patients with mild COVID-19 at day 10^th^ of the illness onset. We also analyzed the relationship of the viral load in the NP samples with the presence/absence of viable particles in cell culture.

## Material and Methods

This study was part of an umbrella project developed in Araraquara, a medium size city from São Paulo State (Brazil) that broadly evaluated the household transmission of SARS-COV-2. The project was approved by the Ethical committee from the Medical School of the University of Sao Paulo (CAPPESQ # 4.235.245). For this particular study, 53 patients with confirmed mild COVID-19 were invited to participate providing nasopharyngeal (NP) samples at the day 10 if illness (D10). Real time PCR using the Altona RealStar® SARS-CoV-2 RT-PCR Kit 1.0 (Altona Diagnostics GmbH, Hamburg, Germany) was run in all samples to confirm the virus presence. The Altona Real Star detects SARS-COV-2 through probes for β-Coronaviruses (targeting gene E) and SARS-COV-2 specific (targeting gene S). For further estimates, we considered gene S since it is the specific for SARS-COV-2.

Samples were stored at −80°C for a maximum of two weeks before being processed for cell culture experiments. Since RT-PCR results were obtained in <24h, we were able to submit some fresh samples to viral isolation (prior the storage at −80°C). From a subset of the positive ones, 0.2ml of the raw samples were put in Vero E6 cells for isolation. We cultured Vero E6 in Dulbecco minimal essential medium (DMEM) supplemented with 10% of fetal bovine serum (FBS), reduced to 2% for the isolation experiments. Two consecutive passages were performed to assure that isolated viruses were replication competent. Positive viral cultures were defined through the observation of cytopathic effect (CPE) in the second passage and increased C_T_ values in the second passage in comparison to the first. Statistical analysis was performed with GraphPad Prism version 7.0. Between-group comparisons were performed using a Student t or Mann-Whitney test.

## Results

Twenty-three of the 53 recruited patients were female (43%) and the median age was 39 years (range 17-60). Forty from the 53 patients were SARS-COV-2 positive at D10 (79%) with the median C_T_ in S gene of 25.7 (range 12-32). From these, 29 had sufficient well-preserved material and were submitted to viral isolation in Vero cell culture.

After two cell-passages, eight samples presented cytophatic effect and positive RT-PCR in Vero cells and 21 did not. However, only seven (24%) filled the criteria CPE plus lower Ct than the original sample in the second passage. Three of the 8 patients with visible CPE at D10 had NP sample also collected at D13, and the material was submitted to virus isolation. However, no RCV was recovered at this time (Table 1).

**Table1.** Information regarding the virus isolation of samples at the 10^th^ day of illness.

| ID | Sample C<sub>T</sub> at D10 | Viral isolation at D10 | C<sub>T</sub> in the cell culture | Presence of Symptoms | Viral isolation at D14 |
| --- | --- | --- | --- | --- | --- |
| 86057 | 28/27 | Negative |  | Nd |  |
| 86059 | 13/12 | **Positive** | 12/10 | No |  |
| 86077 | 21/20 | **Positive** | 13/12 | No |  |
| 86079 | 30/29 | Negative | - | Yes |  |
| 86080 | 31/30 | Negative | - | No |  |
| 86087 | 34/36 | Negative | - | No |  |
| 86088¶ | 22/21 | **Positive** | 25/24 | Nd | Negative* |
| 86089 | 32/31 | Negative | - | No |  |
| 86091 | 32/31 | Negative | - | Yes |  |
| 86109 | 28/27 | Negative | - | Nd |  |
| 86113 | 21/20 | **Positive** | 12/11 | Nd | Negative** |
| 86128 | 20/19 | **Positive** | 11/10 | Nd | Negative*** |
| 86129 | 26/25 | Negative | - | Nd |  |
| 86195 | 20/19 | **Positive** | 10/9 | Yes |  |
| 86196 | 22/21 | **Positive** | 9/8 | Yes |  |
| 86224 | 30/29 | Negative | - | Yes |  |
| 86236 | 28/27 | Negative | - | Yes |  |
| 86237 | 28/28 | Negative | - | Yes |  |
| 86239 | 28/28 | Negative | - | Nd |  |
| 86263 | 28/28 | Negative | - | Yes |  |
| 86364 | 34/34 | Negative | - | Yes |  |
| 86614 | 32/31 | Negative | - | Yes |  |
| 86705 | 28/27 | Negative | - | Nd |  |
| 86708 | 27/26 | Negative | - | No |  |
| 86748 | 25/24 | Negative | - | Yes |  |
| 86750 | 34/32 | Negative | - | Yes |  |
| 86812 | 33/30 | Negative | - | No |  |
| 86815 | 21/19 | **Positive** | 11/10 | Yes |  |
| 86816 | 25/23 | Negative | - | No |  |

Note. The C<sub>T</sub> values are presented for the gene E /gene S.
¶ - Samples submitted with CPE but Ct in cell culture higher than the original sample.
C<sub>T</sub> of the NP samples at D14 were respectively: \* 27/28; \*\*28/27 and \*\*\*27/26

Regarding the symptoms, three of the seven patients with confirmed RCV presented some symptom at the D10, though all of them reported that the symptoms were milder than those experienced during the acute phase (i.e. headache, loss of smell and tiredness – data not shown). It is noted that two patients with viable particles reported no symptoms at all at D10 (Table 1), and two patients did not answer the question.

Among patients from whom no RPV was recovered (22 samples), 7 reported symptoms at D10, nine reported no symptom at this time and 6 did not answer the question. When we looked at the viral load in NP samples, we found that positive samples at cultures presented significant lower C_T_ (20-IQR, 16.5-21) in comparison to negative samples at culture (C_T_29 -IQR, 24-32.2), *p*<0.0001 (Table 1 and Figure 1).

**Figure 1.**
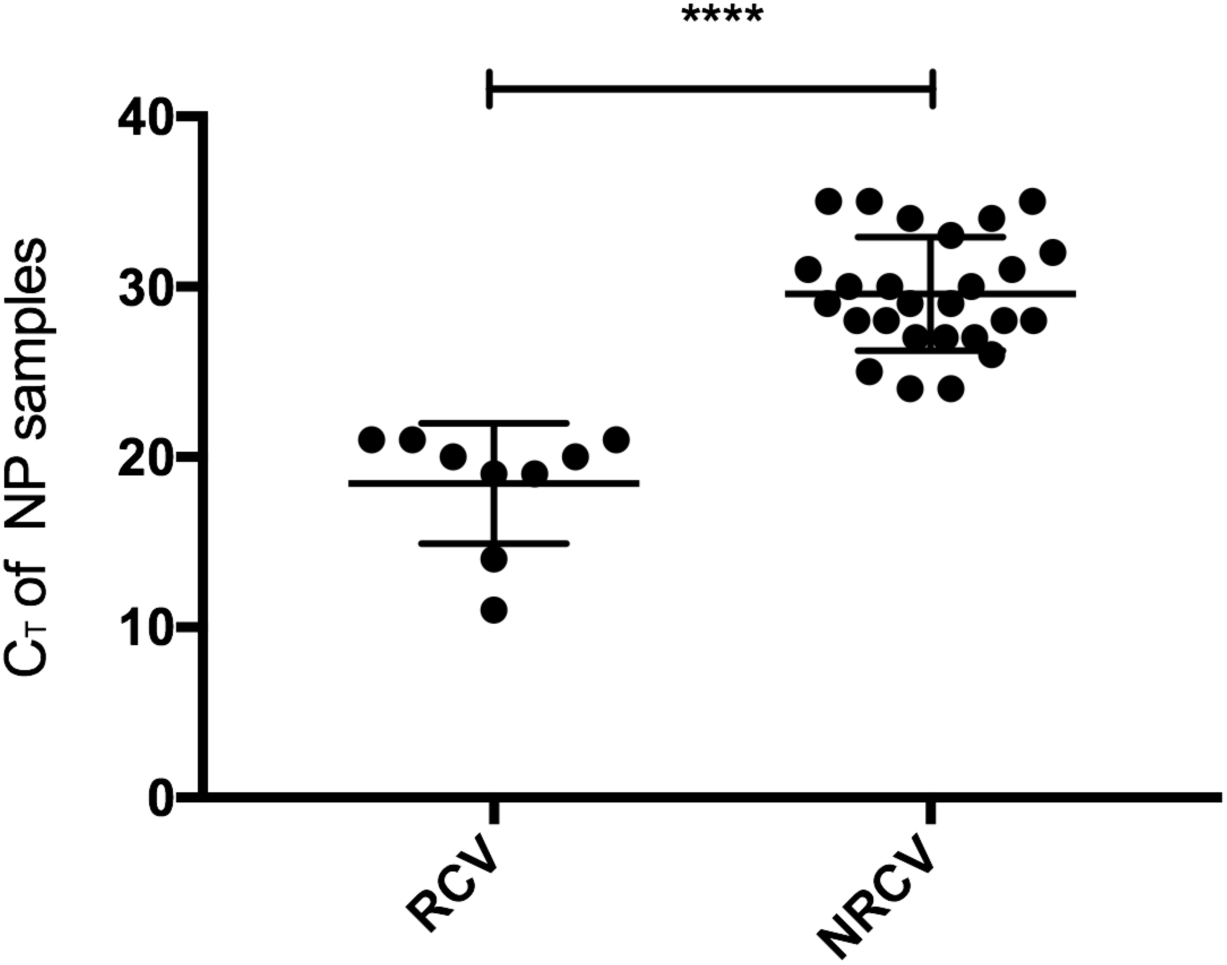
Plot of cycle threshold (C_T_) of the clinical samples at Day 10 in regard to the presence of replication competent virus (RCV) or not (NRCV) in cell culture experiment. Significance was p< 0.0001.

## Discussion

The primary recommendation of 14 days isolation for COVID-19 patients was based on description that individuals with moderate/severe disease could remain RT-PCR positive until day 12-14 [2,3,8,10]. A comprehensive meta-analysis emphasizes the need of isolating the patient early, since the most infectious period is during the first week after symptoms onset. The authors also noticed that no viable particles were detected after nine days from the symptoms onset despite positive RT-PCR in the clinical samples [3]. Based on these evidences, the suggested time for discharge the COVID-19 patient from isolation was reduced to 10 days considering the impossibility to perform molecular tests and the absence of symptoms at the time. Unfortunately, only three studies carefully evaluated the positivity in cell culture along the days [5-7].

The number of studies using cell system to determine the presence of viable SARS-COV-2 particles in clinical samples is still exceedingly small. Among the reasons, there are the requirement of special laboratory facilities and availability of well-conserved samples. For diagnostic purposes, viral culture is also in disadvantage in comparison to RT-PCR, especially during an epidemic situation. Yet, the presence of infectious viruses in cultured cells is the surrogate marker of infectivity.

We report here that mild COVID-19 patients can still be contagious at D10 from the illness onset even in the absence of symptoms. Positivity in cell culture was also strongly associated to cycle thresholds lower than 21 in S gene at the NP samples.

As already observed by others [6], the presence of replication-competent virus was dependent on the viral load in the original sample. However, the authors associated the presence of RCV to both C_T_ and the time interval from disease onset, since no patient sampled after D8 presented viable viruses. In fact, our data agrees to the link between C_T_ values of the sample and SARS-COV-2 growth in cell culture but it confronts the current recommended approach of 10 days isolation of COVID-19 patients as a safe measure to prevent transmission. More important, we were able to recover viable viruses from two patients even after the resolution of symptoms, indicating that this parameter may be not reliable to determine who may or may not be discharged from isolation.

It has to be mentioned that NP sample from one COVID-19 patient presented no increase in viral load at the second passage in cell culture in comparison to the original NP sample (ID #86088), though CPE was observed. It is possible that only a few particles of the original sample were actually replication-competent and therefore, two passages were not sufficient to observe a significant increase in the viral load. Thus, we should be hesitant to state that this particular sample was actually infectious at D10. Limitations of this study include the limited volume of clinical sample that precluded the viral isolation of all positive samples in D10 and the absence of successive clinical samples that would allow us to determine how long the individuals were still infectious.

In summary, available data on SARS-COV-2 culture along the disease are scarce, but indicated so far that persons with mild or moderate COVID-19 remain infectious no longer than 9 days after the symptom onset. Here we present for the first time evidences that individuals with C_T_ values <21 in clinical samples at the 10^th^ day present replication competent viruses at this time. It is crucial to have in mind that we are not fully aware about the sensitivity of the viral culture to recover SARS-COV-2 viable particles, which means that negative viral cultures do not necessarily implies that no infectious particle is present at all. Further studies are needed to determine if viable viruses can occur, as well as in what proportion, between the 10^th^ and 14^th^ days after disease onset. Transmission control strategies focused solely on the presence/absence of symptoms and qualitative PCR results to determine isolation time may be not sufficient to prevent SARS-COV-2 transmission. This work has important implications on the public health and epidemiological surveillance focused on mitigate the SARS-COV-2 transmission chain.

## Data Availability

no new data was generated in this work. Detailed information regarding the results are available upon request.

